# Community Perspectives on PCB Exposure, Health Risks, and Exposure-Reduction Strategies in Guánica, Puerto Rico, Home to a Newly Designated Superfund Site

**DOI:** 10.64898/2026.09.23.26363578

**Authors:** Nicolle Acuña, Johana Ortiz, Noelia Estévez De Rosario, Sara St. George, Samantha González, Olveen Carrasquillo, Laura T Domenech, Ana Ortiz, Nancy Coredor-Cardona, Luis Anthony Alvarez Cruz, Saned Ramos Ramos, Michael Gabriel Silva, Jose A. Rodriguez, Naresh Kumar

**Affiliations:** Department of Public Health Sciences, University of Miami Miller School of Medicine, Miami, FL 33136, USA; Department of Internal Medicine, University of Miami Miller School of Medicine, UHealth, Miami, FL 33136, USA; Senior Medical Officer, Ponce Medical School Foundation; University of Puerto Rico Comprehensive Cancer Center, San Juan, PR; Guánica Students and PR-SPIRIT, Ambassadors, Guánica Puerto Rico

**Keywords:** polychlorinated biphenyls (PCBs), Puerto Rico, environmental justice, qualitative methods, risk perception, community engagement, community health workers

## Abstract

**Background:** Guánica, Puerto Rico, has one of the highest concentrations of environmental polychlorinated biphenyls (PCBs). This PCB contamination coupled with persistent poverty, population loss, and repeated environmental disasters has taken a heavier toll on Guánica community. Our environmental sampling has documented high levels of PCBs in sediment, air and fish, but far less is known about how residents themselves understand exposure, interpret health risk, and weigh contamination against the other pressures of daily life.

**Methods:** We used a qualitative descriptive design nested within the larger HELP Study. This paper presents the final analysis of all qualitative data collected in Guánica: ten sessions comprising five focus groups and five interviews with fishermen (N = 31), conducted between November 2024 and October 2025 in familiar community settings. Sessions followed a semi-structured guide translated from English to Spanish and back-translated for accuracy. Recordings were transcribed in Spanish, verified against the audio, and analyzed in NVivo using codebook-style thematic analysis; three bilingual researchers coded collaboratively and resolved disagreements by consensus.

**Results:** Five themes emerged from our analysis concerning how residents and fishermen understand and navigate PCB contamination: (1) perceived community decline shapes how residents prioritize environmental risks; (2) limited knowledge, misinformation, and communication gaps affect understanding of PCB contamination; (3) cancer concerns are high, but links to PCB exposure remain unclear; (4) PCB contamination is seen as a threat to livelihoods, especially fishing and tourism; and (5) deep mistrust and a desire for accountability influence perceptions of environmental interventions. Participants described contamination not as an isolated hazard but as one strand of a longer history of economic decline, disaster, and institutional neglect.

**Conclusions:** Social, economic, and environmental conditions together shape how Guánica residents perceive contamination of their environment. Meaningful progress requires sustained community involvement, support for local initiatives, development of a Community Health Worker program, and expanded partnerships with local institutions and schools to close communication gaps and promote long-term resilience to address the health effects of PCB contamination in Guánica.

## 1. INTRODUCTION

Polychlorinated biphenyls (PCBs) are synthetic “forever chemicals” that persist in the environment and continue to affect human health long after their use was banned. Their chemical stability allows them to resist natural breakdown and accumulate in soil, water, and living organisms, which makes them a lingering concern for communities exposed through contaminated ecosystems ^2^. Throughout the early twentieth century, PCBs were widely used in industrial processes such as transformers and capacitors, and improper disposal following the 1970s ban contributed to widespread contamination that remains present today ^3^.

Guánica, Puerto Rico, is one of the most affected sites in the world, with PCB levels reported among the second highest globally due to runoff from operations such as Ochoa Fertilizer Company and González Chemical Industries.^4^ Soil on the eastern side of the bay has measured 465 ppm of PCBs, air near the bay 940 pg/m³ against 138 pg/m³ at a background site in Yauco, and fish heads taken from the bay 11,465 ng/g against 859.5 ng/g in fish caught at the bay’s entrance ^5^. PCBs are also not the only contaminant of concern, as state, federal, and university sampling has identified toxic metals, polyaromatic hydrocarbons, and organochlorine pesticides draining from the site into the bay and the low-lying neighborhoods nearby, exposing residents through ingestion, inhalation, and skin contact ^5^. Located on the southwestern coast of the island, Guánica was once known for its sugar mill and industrial factories. Today, it relies heavily on tourism and fishing supported by its beaches and coastal waters. Despite these natural assets, Guánica remains a vulnerable community marked by persistent poverty, an aging and declining population, and some of the highest cancer rates in Puerto Rico ^6,7^. Since receiving a Superfund designation in 2022, it has become increasingly clear that residents are experiencing adverse health outcomes linked to long-term PCB exposure, making this an urgent environmental justice issue ^8^. Figure 1 summarizes the major environmental and management factors contributing to PCB contamination in Guánica Bay, including origins, transport mechanisms, exposure pathways, and community impacts.

**Figure 1.**
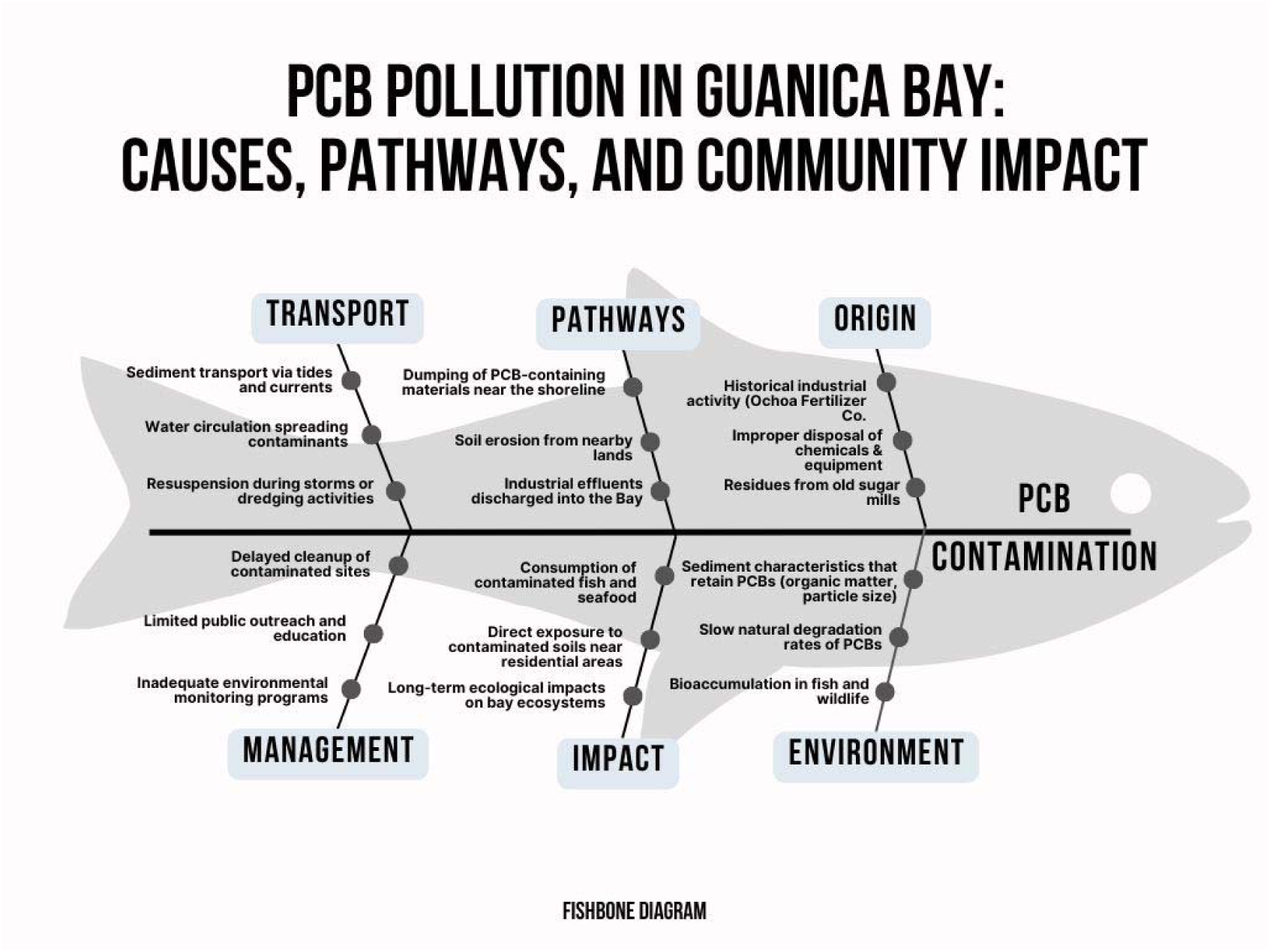
Fishbone diagram illustrating the origins, transport mechanisms, exposure pathways, environmental factors, and community impacts contributing to PCB contamination in Guánica Bay.

The health risks associated with PCBs are well documented. Exposure contributes to oxidative stress and chronic inflammation, which increase the risk of breast, prostate, and liver cancers and are linked to cardiovascular and metabolic diseases ^9^. Children face particularly serious risks, as PCBs accumulate in the body over time and interfere with brain development, leading to cognitive impairments, learning difficulties, and behavioral challenges ^10^. These impacts intensify existing health disparities and add new barriers for residents seeking preventive care or reliable information ^11^.

Fishermen are an especially important subgroup within this context because they are directly exposed through daily contact with contaminated water and through frequent consumption of local fish. Their livelihood depends on the bay, so contamination not only poses health risks but also threatens the economic stability of families who rely on fishing as their primary source of income. Understanding their experiences helps clarify how contamination affects both exposure pathways and community wellbeing.

At the same time, Guánica’s residents must navigate these risks while also facing limited access to clear, consistent health communication. Many community members rely on informal networks, personal experience, or social media to understand contamination, which can contribute to confusion or mistrust ^8^. Addressing PCB exposure requires communication strategies that are culturally grounded, accessible, and developed in collaboration with the community ^12^. Exposure reduction practices, such as avoiding contaminated areas or preparing fish by removing fatty tissue and baking, remain important tools for lowering PCB intake without increasing fear or stigma ^13,14^.

Although previous environmental sampling research has established the presence and severity of PCB contamination, these data cannot describe how residents interpret risk, how contamination affects their decisions, or what barriers shape their ability to respond. Qualitative work is needed to understand lived experiences, gaps in communication, and the broader social and economic pressures that influence how people make sense of environmental hazards.

Understanding how residents interpret PCB risks, what information they receive, and how contamination shapes their daily lives is essential for designing meaningful public health interventions. This study used a codebook-style thematic analysis to center community perspectives on PCB contamination in Guánica and to inform community-driven public health strategies.

## 2. MATERIALS AND METHODS

### 2.1. Participants and Setting

Guánica is a small coastal community whose daily life and local economy depend closely on its waters, beaches, and fishing activity. Because residents’ routines are so closely tied to their environment, it was essential to collect information directly within the community to understand how people interact with the spaces affected by contamination. All focus groups and interviews were conducted in person during field visits to Guánica at locations already familiar to participants, including churches, schools, community centers, and fishermen’s shops. Holding sessions in these everyday community settings allowed participants to speak openly and fostered a sense of comfort, trust, and partnership between researchers and residents.

Eligible participants included adults 18 years and older who were residents of Guánica and/or local fishermen. Participants were recruited through community outreach in public locations such as community centers, churches, and fishermen’s shops. A community coalition of four Guánica residents supported recruitment by distributing flyers and engaging with community members about the study, helping to maintain the community’s trust in preparation for the arrival of the Miami-based research team ^12^. Residents who were informed about the study and interested voluntarily attended the interviews or focus groups.

This work is part of the larger HELP Study ^4^, which includes approximately 200 participants. This paper presents the final qualitative analysis of all sessions conducted in Guánica: ten sessions completed between November 2024 and October 2025, comprising five focus groups and five interviews with fishermen, representing 31 residents and fishermen of Guánica and nearby communities. Twenty-six residents took part in the focus groups, and five fishermen took part in the interviews. The focus group participants were mostly women, with a mean age of 49.1 years. The fishermen interviewed were all men who relied on fishing as their primary livelihood, with a mean age of 58.4 years.

### 2.2. Procedure

This study received IRB approval from the University of Miami, and all members of the research team completed CITI training. All interviews and focus groups were conducted by bilingual female researchers from the Department of Public Health Sciences at the University of Miami (N.A., J.O., N.E.R.).

Participants completed a brief sociodemographic survey that our research team developed to capture key characteristics such as age, gender, and health history, including chronic diseases and cancer. We administered the survey to participants in Guánica and in communities across Puerto Rico to better understand the broader context of environmental health in the region. For the purposes of this qualitative study, we do not present the survey findings, as our analysis focuses on the narrative and experiential data gathered through focus groups and interviews; results from the companion quantitative work in Guánica are reported separately ^5^.

The final analysis included five focus groups averaging 57 minutes and five interviews with fishermen averaging 40 minutes. Prior to participation, all individuals provided verbal consent, and interviewers assured participants of confidentiality. The facilitators used a discussion guide to navigate the focus group sessions and create a welcoming environment. This guide was translated from English to Spanish and underwent back translation to ensure accuracy. Questions ranged from life in Guánica to awareness and understanding of contamination, health, life and economic impacts, experiences with regulatory agencies, community voice and participation, and future visions and needs. Table 1 presents the full qualitative interview question guide used in these sessions.

**Table 1.**
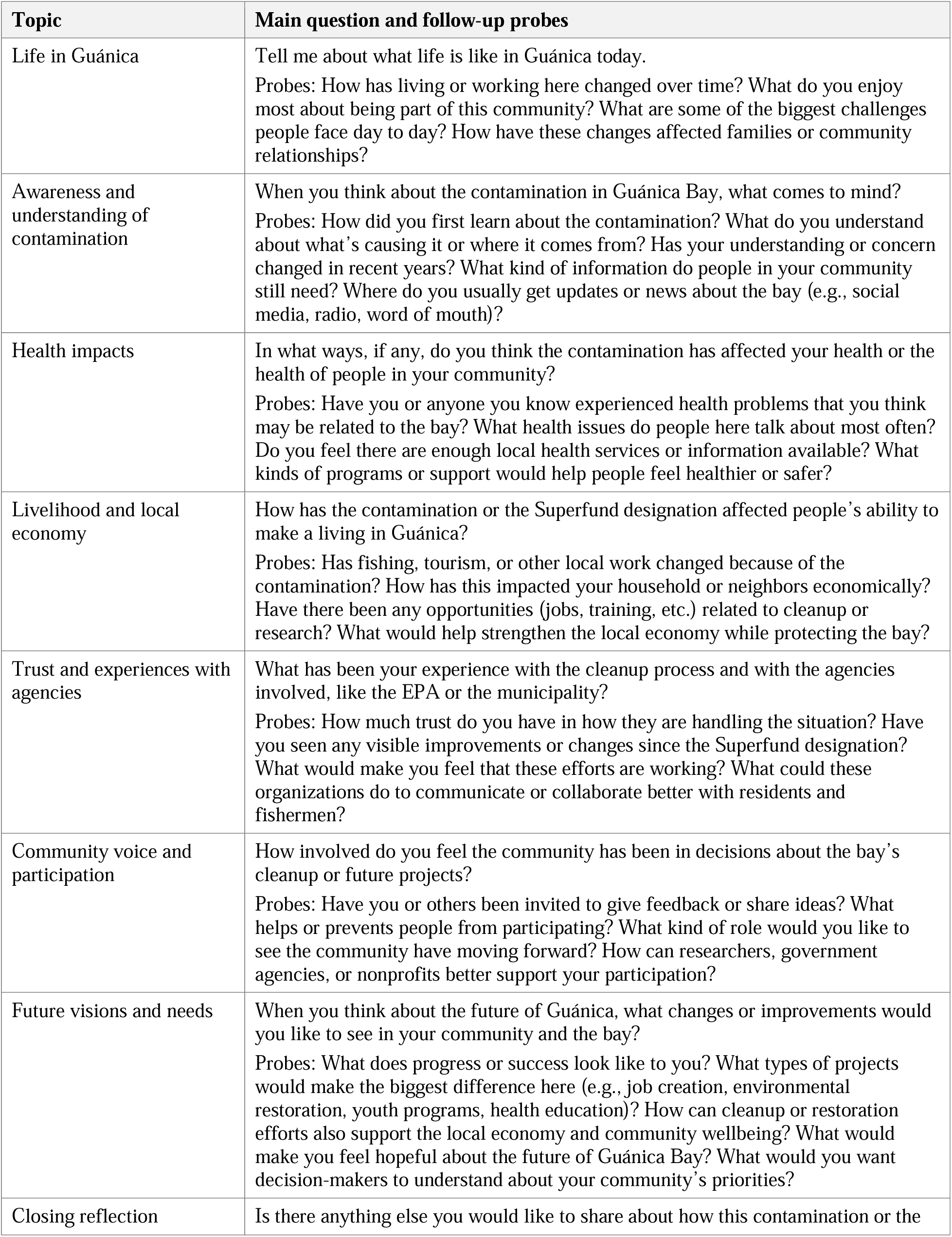
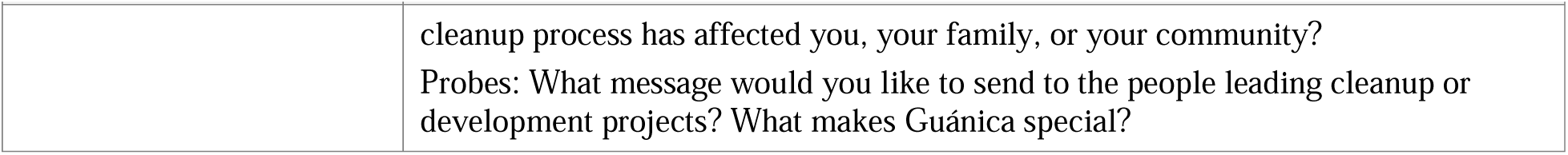
Qualitative interview question guide.

| Topic | Main question and follow-up probes |
| --- | --- |
| Life in Guánica | Tell me about what life is like in Guánica today.<br>Probes: How has living or working here changed over time? What do you enjoy most about being part of this community? What are some of the biggest challenges people face day to day? How have these changes affected families or community relationships? |
| Awareness and understanding of contamination | When you think about the contamination in Guánica Bay, what comes to mind?<br>Probes: How did you first learn about the contamination? What do you understand about what's causing it or where it comes from? Has your understanding or concern changed in recent years? What kind of information do people in your community still need? Where do you usually get updates or news about the bay (e.g., social media, radio, word of mouth)? |
| Health impacts | In what ways, if any, do you think the contamination has affected your health or the health of people in your community?<br>Probes: Have you or anyone you know experienced health problems that you think may be related to the bay? What health issues do people here talk about most often? Do you feel there are enough local health services or information available? What kinds of programs or support would help people feel healthier or safer? |
| Livelihood and local economy | How has the contamination or the Superfund designation affected people's ability to make a living in Guánica?<br>Probes: Has fishing, tourism, or other local work changed because of the contamination? How has this impacted your household or neighbors economically? Have there been any opportunities (jobs, training, etc.) related to cleanup or research? What would help strengthen the local economy while protecting the bay? |
| Trust and experiences with agencies | What has been your experience with the cleanup process and with the agencies involved, like the EPA or the municipality?<br>Probes: How much trust do you have in how they are handling the situation? Have you seen any visible improvements or changes since the Superfund designation? What would make you feel that these efforts are working? What could these organizations do to communicate or collaborate better with residents and fishermen? |
| Community voice and participation | How involved do you feel the community has been in decisions about the bay's cleanup or future projects?<br>Probes: Have you or others been invited to give feedback or share ideas? What helps or prevents people from participating? What kind of role would you like to see the community have moving forward? How can researchers, government agencies, or nonprofits better support your participation? |
| Future visions and needs | When you think about the future of Guánica, what changes or improvements would you like to see in your community and the bay?<br>Probes: What does progress or success look like to you? What types of projects would make the biggest difference here (e.g., job creation, environmental restoration, youth programs, health education)? How can cleanup or restoration efforts also support the local economy and community wellbeing? What would make you feel hopeful about the future of Guánica Bay? What would you want decision-makers to understand about your community's priorities? |
| Closing reflection | Is there anything else you would like to share about how this contamination or the |
|  | cleanup process has affected you, your family, or your community?<br>Probes: What message would you like to send to the people leading cleanup or development projects? What makes Guánica special? |

Focus groups and interviews were audiotaped, and participants provided verbal consent prior to participating. Interviews were not repeated after initial discussions, as participants covered all questions during the allotted time. Audio recordings from the focus groups and interviews were transcribed in Spanish by N.A. To ensure accuracy, J.O. and N.E.R. reviewed each transcript while simultaneously listening to the audio, verifying that participants’ responses were captured correctly. To maintain participant privacy, all names and identifying information were removed from the transcripts so that comments shared during the focus groups and interviews remained anonymous. Transcripts were not returned to participants for comments or corrections due to the study’s timeline.

### 2.3. Data Analysis

We analyzed the data using a codebook-style thematic analysis, consistent with Braun and Clarke’s description of thematic analysis as a flexible family of approaches that includes structured, collaboratively developed codebooks ^15,16^. N.A. drafted the initial codebook after a preliminary review of transcripts and field notes. The three coders (N.A., J.O., N.E.R.) then refined the codebook collaboratively during early coding sessions, revising definitions, adding inductive codes, and reorganizing categories to reflect concepts that emerged directly from participants’ narratives. The full codebook, including code definitions and example excerpts, is presented in Appendix Table A1.

We coded the transcripts together in real time using NVivo software, referencing field notes to contextualize participant statements, clarify meaning, and support the development of new codes. Field notes were especially important for capturing nonverbal cues, community dynamics, and environmental context observed during data collection in Guánica. After coding each transcript, we discussed areas of agreement and disagreement until reaching consensus before moving to the next transcript.

Throughout the process, we consulted with a faculty member who specializes in qualitative methods to ensure analytic rigor. A content expert (N.K.) provided guidance on environmental health concepts and the foundations of the HELP Study ^4^.

### 2.4. Generative AI Disclosure

Generative artificial intelligence was used only for assistance with grammar, organization, and refinement of manuscript text. No transcripts, participant data, or identifiable information were uploaded to any AI platform. Study design, data collection, coding, and analysis were conducted entirely by the research team.

## 3. RESULTS

The study team identified five themes that describe how residents and fishermen in Guánica understand and navigate PCB contamination in Guánica: (1) perceived community decline shapes how residents prioritize environmental risks; (2) limited knowledge, misinformation, and communication gaps affect understanding of PCB contamination; (3) cancer concerns remain high without a clear link to PCB exposure; (4) PCB contamination threatens livelihoods, especially fishing and tourism; (5) deep mistrust and a desire for accountability influence perceptions of environmental interventions. These themes show how local experiences, social conditions, and environmental realities inform the community’s interpretations of contamination and its consequences. Additional supporting quotes for each theme are presented in Appendix Table A2.

### 3.1. Perceived Community Decline Shapes How Residents Prioritize Environmental Risks

Participants described PCB contamination within the broader context of longstanding economic decline, population loss, and the cumulative effects of hurricanes, earthquakes, and other environmental disasters. These overlapping stressors were frequently mentioned when discussing with community members. For example, one resident noted, “I think there are multiple, many variables at play, and they’re all acting together, you can’t just say it’s pollution” (Participant 3, Focus Group 2).

Residents explained that the community has endured repeated crises, which has shaped how they view the urgency of contamination. Several noted that concerns about PCB exposure were often secondary to more immediate financial and structural challenges:

There’s only one fact. Guánica is the poorest town in Puerto Rico, we are the Cinderella. So what do you think…has it ever really mattered much? They contaminated us. But there was work, just like at the sugar mill…but there was work. (Participant 1, Focus Group 1)

Participants also described how natural disasters and the Superfund designation have further contributed to population decline, with some residents perceiving the town as abandoned or unlikely to recover. They noted that the contamination, combined with these disasters, has led some individuals to view Guánica as a “lost cause,” discouraging former residents from returning. One participant talked about people “…leaving Guánica because if there’s nothing here, people will just keep leaving the town” (Fisherman, Interview 1).

### 3.2. Limited Knowledge, Misinformation, and Communication Gaps Affect Understanding of PCB Contamination

Community members had general awareness of PCB contamination but lacked clear, accurate knowledge about PCBs, their sources, and health effects. Heavy reliance on word of mouth, social media, and informal networks contributed to the spread of information concerning the contamination of the bay. For example, one participant recalled, “Ever since I was little, my mom always told me that you couldn’t go into the water by the pier” (Participant 3, Focus Group 2).

Participants frequently described feeling excluded from decision-making processes and inadequately informed by government agencies or outside organizations. This lack of direct communication contributed to uncertainty about how to protect themselves. One participant noted, “No one tells us exactly what it means for us or how to protect ourselves” (Participant 2, Focus Group 1).

Residents also described how information about the contamination is often distorted or exaggerated as it spreads through the community. For example, some referenced claims that “the National Oceanography Organization found contaminant particles 3,000 miles from the bay” (Participant 4, Focus Group 1).

### 3.3. Cancer Concerns Are High, but Links to PCB Exposure Remain Unclear

Participants frequently discussed cancer and chronic illness within their families and broader social networks. Many described hearing about neighbors, relatives, or community members who had become ill, and several felt that cancer had become increasingly common in Guánica. This concern emerged in comments such as:

[I’m in my 70s.] I’ve never suffered from anything. What I do think is that before, I understood that cancer ran in families — you know, like something you try to avoid. And now, all of a sudden, everyone has cancer…In my family there are cases of cancer, and of different types of cancer. (Participant 5, Focus Group 1)

While some participants believed that contamination might be related to these health patterns, others did not make a connection or expressed uncertainty about any link to PCB exposure. Residents referenced a variety of explanations circulating in the community, including concerns about “dirty water” and “poisonous fish.”

### 3.4. PCB Contamination Is Seen as a Threat to Livelihoods, Especially Fishing and Tourism

Participants described PCB contamination as directly affecting key sources of income in Guánica, particularly fishing and tourism. Fishing was described as a primary livelihood for many residents, and participants noted that the contamination had altered community perceptions of seafood safety. Fishermen reported receiving significant backlash from community members who feared consuming contaminated fish. Several described customers refusing to purchase seafood and questioning the safety of what was being sold:

I got so many calls and messages, so many, really so many people. ‘I bought fish from you, didn’t I? Did you know that fish wasn’t good, that it was poisoned? I’m not buying fish from you anymore…Why don’t you all put up a sign saying the fish is poisoned?’ That was people’s attitude, because most of them don’t understand, you know, they didn’t understand. (Fisherman, Interview 1)

Some participants reported that they no longer trusted local fish at all, with statements such as “Now, I’m dying to eat a fish and I don’t dare. Now, I crave for fish, but I don’t try it” (Participant 7, Focus Group 1). Participants described how this fear extended beyond fishermen to affect local restaurants and businesses along the bay, as residents and visitors sought to avoid perceived risks.

### 3.5. Deep Mistrust and a Desire for Accountability Influence Perceptions of Environmental Interventions

Participants described limited trust in government agencies, industries, and external organizations involved in addressing PCB contamination. Many felt that communication from the Environmental Protection Agency (EPA) and local authorities lacked transparency, and some believed that important details about the contamination or cleanup process were being minimized or withheld. As one resident put it, “You can’t put goats in charge of lettuce” (Participant 2, Focus Group 2).

Several participants also criticized industries associated with the contamination, including Ochoa Fertilizer Company, describing them as prioritizing their own interests rather than the wellbeing of the community. One resident expressed this frustration:

And the negligence of the authorities in this regard is worrying. The struggle is immense, and sometimes lies and other factors outweigh the truth. We must continue educating and we must continue fighting. (Participant 3, Focus Group 2)

Participants further described uncertainty about the intentions of outside groups conducting research or community outreach. Some entered the study with skepticism toward external involvement, stating, “At first, I didn’t trust any of this, because sometimes you think it’s just another outsider coming in who’s going to do something and then doesn’t do anything” (Fisherman, Interview 2).

Even with this skepticism, many emphasized the importance of local leadership and expressed interest in having community-guided structures to support advocacy and communication. One participant noted, “I know that if you guys say, like, ‘Oh, let’s do a workshop or a seminar,’ we’ll explain the information, and then we’d be grateful if you spread the word. That would work” (Participant 7, Focus Group 2).

## 4. DISCUSSION

The goal of this study was to understand how Guánica residents and local fishermen interpret PCB contamination, how they make sense of their health risks, and how these concerns fit within the wider challenges their community faces. Our research questions focused on how residents understand exposure pathways, what sources of information they rely on, and how contamination shapes their daily decisions and wellbeing. Looking across all themes, residents did not experience contamination as a single issue. Instead, they connected it to economic decline, repeated natural disasters, mistrust of institutions, and an ongoing struggle to access clear information.

Residents frequently discussed PCB contamination alongside economic hardship, hurricanes, earthquakes, and population loss. This framing aligns with other environmental justice studies, such as research in Texas colonias showing that people interpret environmental hazards through the broader pressures of daily life ^8,12,17^. Guánica adds an important dimension, as repeated disasters have shaped routines and priorities for decades. A major gap in the PCB literature is that most studies focus on chemical exposure rather than the lived experience of communities navigating long-term decline and recovery ^4^. Our findings show how this context influences how urgent PCB risks feel and why some residents focus more on immediate concerns than long-term exposures.

Limited knowledge, misinformation, and inconsistent communication also shaped uncertainty about contamination. Many residents relied on family, neighbors, or social media for information, which often led to confusion. This pattern mirrors findings from Texas colonias ^12^ and New Bedford Harbor, where fishing advisories were not well understood and culturally relevant communication was limited ^14^. In Guánica, misinformation filled gaps when official communication was unclear or inconsistent. Residents said they had few opportunities to ask questions or receive direct explanations. Very little research exists on PCB communication in Spanish-speaking Caribbean communities, leaving significant gaps in understanding how information spreads across cultural and linguistic contexts.

Residents expressed concern about what they perceived as a high number of cancer cases among family and community members. Fishermen shared stories of friends and colleagues diagnosed with cancer or who died suddenly from unknown causes. Although many grew up hearing that cancer was hereditary, they began questioning this idea when neighbors without a family history became ill. This aligns with findings from other contaminated or rural areas where people rely on illness narratives to interpret environmental risk ^8,12,16^. Yet most residents were unsure how PCB exposure could contribute to these patterns. Concepts such as bioaccumulation of PCBs were minimally understood, making it difficult to follow practices such as preparing fish correctly or avoiding contaminated areas. Scientific studies have shown clear evidence that PCB exposure increases cancer risk ^9,10,13^, and these risks extend beyond cancer to include other chronic conditions and harms to children and developing fetuses. Pregnant women, young children, and older adults would especially benefit from clearer explanations of exposure pathways and guidance on how to reduce risk.

Quantitative work by our group in the same community supports these accounts. In a survey of 419 Guánica residents and 848 residents from four other areas of Puerto Rico, chronic conditions were more common in Guánica, including cancer (3.6% versus 2.5%), cardiovascular disease (18.9% versus 9.7%), and endocrine disorders (29.1% versus 18.3%). Endocrine disorders also tracked with PCB body burden, where a 1% increase was associated with 41% higher odds after adjustment for age, gender, smoking, and alcohol use (OR = 1.41; 95% CI 0.99–2.03). Cancer registry data show a similar pattern, with age-standardized breast and corpus uterus incidence in zip code 00653 rising from 111.4 in 2010–2016 to 168.4 in 2017–2023, while the rate for Puerto Rico overall was significantly lower in the later period (SRR = 0.75; 95% CI 0.59–0.98) ^5^. These comparisons are correlational and cannot prove casual mechanism of diseases due to PCB exposure. However, they suggest that residents who sense an unusual amount of illness around them are reading their surroundings accurately, and that the gap lies in explanation rather than in perception.

Mistrust also shaped how residents interpreted contamination and response efforts. Participants described feeling excluded from decision making, uncertain about the information provided, and concerned that important details were being withheld. Many questioned whether the response would have been different if the contamination had occurred in a wealthier or more visible town. These concerns reflect findings in other environmental justice settings where communities question institutional intentions and feel disconnected from formal risk communication ^8,17^. Although residents appreciated when outside groups communicated clearly and respected local knowledge, many felt that long-term responsibility should shift toward the community itself. Residents expressed a desire to protect one another, rebuild their town, and design programs grounded in their own experiences. This highlights a gap in the literature, as most documented interventions in Caribbean settings focus on short-term educational campaigns rather than long-term, community-led strategies ^8,18^.

Participants described how contamination could harm local fishing and tourism, which are central to Guánica’s economy and identity. This aligns with research showing that environmental contamination affects income, cultural traditions, and the sense of place linked to generational fishing practices ^14^. Fishermen want to protect their health, yet they depend on fishing to sustain themselves and maintain the town’s cultural history. Limited research exists on how contamination affects the emotional and economic experiences of fishermen in Puerto Rico, and our findings point to a need for more work in this area. Any intervention in Guánica must incorporate fishermen directly, since the goal is to support their work rather than harm their business. Many fishermen already understand which areas should not be fished, yet described challenges convincing customers that their catch is safe. This suggests an opportunity for a local certification process that could reassure customers and protect income. The literature also provides little insight into how a Superfund designation affects long-term economic stability or community recovery. Residents and fishermen described how contamination influences how outsiders view Guánica and how local businesses struggle with negative assumptions. These perceptions must be considered in future recovery planning.

### 4.1. Implications and Future Directions

Looking across all the themes, it appears that Guánica needs communication that is simple, bilingual, and consistent, and that reaches people where they already gather. Most residents learn about contamination through family and neighbors, which shows how much room there is to improve the way information is shared ^12,19^. We partnered with the local high school and science teachers to involve students in an environmental poster competition while teaching them about Guánica Bay. Continuing the poster competition will help strengthen this effort by educating students about contamination in their community, encouraging them to take action, and helping them share accurate information at home. Since students are the future of Guánica, investing in their learning can support long-term change. Moving forward, communication should stay honest, easy to understand, and focused on building lasting relationships with the community, not just offering information once ^8^.

Residents also want to be more involved in decisions and planning. They want the chance to ask questions, understand what is happening around them, and help shape the solutions. This is similar to what other studies have found in communities facing environmental issues. When people are involved from the beginning, they feel a stronger sense of ownership and trust ^8,19^. A Community Health Worker (CHW) program or a local community coalition could help meet this need by providing a reliable and trusted presence in Guánica. These programs can reduce misinformation, support families, and create a stronger link between researchers, agencies, and residents ^19^.

Another important consideration is how contamination affects people’s day-to-day lives, especially because Guánica has such a long history of economic challenges and natural disasters. Interventions that do not consider these realities are less likely to be useful or respectful ^7,17^. The community’s everyday experiences must guide the next steps.

We also learned several practical lessons. Building trust takes time and should start well before data collection, along with good organization and crowd management to make group discussions run more smoothly. Understanding the broader community context is key when working in rural areas like Guánica, as environmental concerns can be closely tied to local politics and long-standing frustrations ^15,19^. As our work continues, we plan to expand environmental health education in local schools, especially with high school students, and begin developing a CHW program that can offer ongoing support. We also want to keep listening to residents and fishermen so that future efforts reflect what matters most to them. Strengthening partnerships with community groups, Ponce Health Sciences University, and local organizations will be essential.

Overall, addressing PCB contamination in Guánica requires more than laboratory testing or cleanup plans. It requires listening to residents, supporting their leadership, and creating systems that make space for their voices. The community is resilient, resourceful, and willing to work toward a healthier future. Both residents and fishermen shared their hopes for Guánica’s future. Many talked about wanting the town to become a place people from all over would want to visit again. They expressed pride in their beaches, landscapes, and cultural history, and described a strong desire to restore areas like the Malecón so they can be places where the community and visitors gather. Their vision for the future is rooted in safety, beauty, and resilience. Across these conversations, the shared goal was clear: to restore Guánica in a way that honors its identity, protects its people, and supports the livelihoods that have sustained the town for generations.

### 4.2. Strengths and Limitations

This study has several strengths that helped us better understand how Guánica residents and fishermen make sense of PCB contamination. Collecting data in person and in familiar community spaces allowed participants to feel comfortable and speak openly. Coding the data together as a team, using detailed field notes, and working closely with local partners strengthened the quality and depth of the analysis. These approaches helped us capture perspectives that might have been missed through more formal or distant research methods.

There are also limitations that should be acknowledged. Although this analysis includes all focus groups and fishermen interviews conducted in Guánica, participants were recruited through community outreach and may not fully represent the experiences of youth, seasonal residents, or people less connected to fishing or tourism. Although we are bilingual Latino researchers, we are not Puerto Rican and are based in Miami. These differences may have shaped both how participants viewed us and how we interpreted their experiences. Future work should continue to collaborate with local leaders and expand perspectives to include other groups within the community.

## 5. CONCLUSIONS

This study contributes to the Superfund Research Program by demonstrating disproportionate burden of exposure to hazardous substances in communities living around Superfund site. Guánica residents have been experiencing economic decline, repeated disasters, and institutional mistrust. By centering residents’ and fishermen’s voices, we highlight the need for community-driven, culturally grounded strategies that support both environmental remediation and social resilience. Developing a Community Health Worker program, strengthening partnerships with local institutions, and expanding school-and community-based education efforts are promising next steps to close communication gaps and support Guánica residents in shaping the future of their community.

## Abbreviations

The following abbreviations are used in this manuscript
CHW: Community Health Worker
EPA: U.S. Environmental Protection Agency
PCB: Polychlorinated biphenyl

## Author Contributions

Conceptualization, N.A. and N.K.; methodology, N.A.; software, N.A., J.O., and N.E.R.; validation, N.A., J.O., and N.E.R.; formal analysis, N.A., J.O., and N.E.R.; investigation, N.A., J.O., and N.E.R.; resources, N.A., N.K., and S.G.; data curation, N.A., J.O., and N.E.R.; writing—original draft preparation, N.A.; writing—review and editing, N.A., J.O., and N.E.R.; review and feedback: NK, LD, NC, AO, OC, LC, JR, SR, MS; visualization, N.A., J.O., and N.E.R.; supervision, N.K. and S.G.; data collection: NA, NK, LC, JR, SR, MS; project administration, N.A. and N.K.; funding acquisition, N.K. and S.G. All authors have read and agreed to the submitted version of the manuscript.

## Institutional Review Board Statement

The study was conducted in accordance with the Declaration of Helsinki and approved by the Institutional Review Board of the University of Miami (IRB #20190217; approval date 10/13/2025).

## Informed Consent Statement

Verbal informed consent was obtained from all subjects involved in the study.

## Data Availability Statement

The data collected for this study include qualitative transcripts that cannot be shared publicly due to privacy and confidentiality protections for participants. De-identified excerpts are included in this paper. Photographs of study settings and data collection activities have been removed to protect participant confidentiality; readers may contact the corresponding author to request access to these materials.

## Data Availability

All data produced in the present study are available upon reasonable request to the authors

## Acknowledgments

We are grateful to the community leaders and residents of Guánica for their support and warm welcome. Special thanks go to the stakeholders in the community who were contacted and helped with recruitment and community coordination.

## Appendix A

**Table A1.** Qualitative analysis codebook.

| Code | Label | Definition and examples | Sample quotes |
| --- | --- | --- | --- |
| **1.0 Sense of place and community identity** |  |  |  |
| 1.1 | Connection to Guánica | Captures participants' duration of residence in Guánica. | — |
| 1.1.1 | Lifelong residents | Born and raised in Guánica. | "Born and raised here, my whole life in Guánica." (P3, FG1)<br>"My whole life living in Guánica." (P4, FG1)<br>"[In my 70s.] We're from the same crew." (P6, FG1) |
| 1.1.2 | Less-connected or newly settled residents | Participants who are newer to the community, have lived there for many years but feel limited connection or awareness, or left and later returned with weaker community ties. | "I've been living here for [many] years, but before that I just went from work to home." (P2, FG1)<br>"I lived abroad for many years and came back, but I'm not that up to date on everything." (P2, FG1) |
| 1.2 | Community attachment and pride | Expressions of affection, loyalty, or identification with Guánica despite challenges. | "I continue to live happily in my town because I love it." (P5, FG1) |
| 1.3 | Places and activities within the community | Mentions of places or activities that participants engage in within their community. Reflects how residents spend time locally, interact with their surroundings, and participate in community life. | — |
| 1.3.1 | Natural spaces | Beaches, bay, rivers, mangroves. | "I loved the beach; we used to go camping and everything." (P1, FG2) |
| 1.3.2 | Social or communal spaces | Plaza, church, community centers, or public gatherings where people connect and share information. | "At church, they always gave us materials about pollution." (P4, FG1) |
| 1.3.3 | Outdoor activities | Fishing, gardening, walking. | "I like gardening." (P7, FG1)<br>"I walk all the way across town." (P4, FG1)<br>"My daughter has friends whose parents fish." (P2, FG1) |
| **2.0 Environmental concerns** |  |  |  |
| 2.1 | Perceived environmental change | Observations of changes in water, air, or land quality over time. | "The water used to be clean... now it smells bad and looks ugly." (P4, FG1)<br>"That said, the mangroves disappeared, and with them, the fish." (P3, FG2) |
|  |  |  | “Every day the fruit gets worse; it comes out black and it looks like it’s dried out inside. And another thing I don’t understand is that, for example, a soursop turns red inside. I don’t get it.” (P7, FG1) |
| 2.2 | Perceived contamination sources | Mentions of factories, petrochemical industries, or pollution origins (petrochemical plants, Ochoa Fertilizer, wastewater, sargassum). | “When they built the petrochemical plants, the bay was damaged.” (P1, FG1)<br>“They took advantage of the ignorance of government X... they brought in the petrochemical plants.” (P8, FG1)<br>“And on top of that, a ship came, that ended up polluting the area since the petrochemical plants arrived.” (P8, FG1) |
| 2.3 | Environmental neglect and abandonment | Feelings that the area has been ignored or not maintained. | “All of that has been abandoned; nobody cleans the seas.” (P1, FG1)<br>“The sand on the beach where I grew up. People used to lie down, cover themselves, and play in it. Now you can’t even tell what color it is... Now it’s a horrible, dark thing, with a bad smell.” (P8, FG1) |
| **3.0 Health impacts and concerns** |  |  |  |
| 3.1 | Fear or anxiety about contamination | Emotional distress and avoidance behaviors. | “I’m terrified; I don’t even dare to drink water.” (P1, FG1)<br>“I’ll die of something else. Not from the pollution.” (P4, FG1)<br>“Here, we just listen and get scared.” (P6, FG1)<br>“Now, I’m dying to eat a fish and I don’t dare. Now, I crave for fish, but I don’t try it.” (P7, FG1) |
| 3.2 | Perceived health conditions linked to the environment | Health problems residents associate with PCB exposure or other environmental contamination. | “There were eight cases of tuberculosis on the Ochoa highway... it was proven to be due to the severe pollution.” (P4, FG2)<br>“But my sister worked there for many years... she lived there, worked there, she was always there, and always had something. There had to be something there.” (P2, FG1) |
| 3.2.1 | Cancer and chronic illness | Mentions of cancer or long-term disease linked to the environment. | “I’m a lung cancer survivor... I don’t know if it’s the effect of the bay or the petrochemicals.” (P5, FG1)<br>“I can tell you that the incidence of cancer in Guánica is very high. My mother has cancer. I have five friends who are fishermen. Four of them died last year from cancer.” (Fisherman, Interview 2)<br>“And now, all of a sudden, everyone has cancer... In my family there are cases of<br>cancer, and of different types of cancer.” (P5, FG1)<br>“Living and working here, seeing so many health issues, so many people—young people, children, regardless of age—suffering from cancer and other conditions, sometimes you just can’t explain where they come from.” (P1, FG1) |
| 3.2.2 | Respiratory issues and allergies | Asthma, allergies, and breathing problems tied to air or dust. | “That dust can be harmful... especially to those with asthma.” (P7, FG1)<br>“Perhaps the asthma I developed about 15 years ago is due to that.” (P7, FG1)<br>“I have to take an allergy pill, one in the morning and one at night.” (P7, FG1) |
| 3.2.3 | Child and family health concerns | Expressed feelings of anxiety or references to illnesses in children or younger generations. | “A person born to a sick mother... and not just with cancer, but with many autism spectrum disorders.” (P1, FG1)<br>“It was said that many people who swam there, like a little girl who said she came out of Playa Santa with a very delicate condition.” (P4, FG1) |
| 3.3 | Access to health resources and testing | Perceived gaps in medical support, screening, or governmental health response. | “Well, these medical treatments don’t have medical coverage, and nobody cares about them. See?” (P6, FG1)<br>“...we older folks are the ones who die, and we don’t have health insurance...” (P5, FG1) |
| 3.3.1 | Need for testing and monitoring | Residents’ demand for scientific health testing and monitoring. | “They should test our bodies; nobody checks the water or the land.” (P1, FG1) |
| 3.3.2 | Transparency in health research | Distrust or lack of understanding of the health research process. | “So you’re interrogating us. Then we’ll find out what we talked about here, what we did.” (P6, FG1)<br>“What we need to hear is the voice of reason.” (P4, FG1) |
| **4.0 Economic dependence and decline** |  |  |  |
| 4.1 | Loss of livelihood | Loss of economic stability in Guánica, among residents whose general job opportunities have diminished and among those whose income once depended directly on the bay. | “There are no more sugar canes... Now, all there is is cement... We are the poorest town in Puerto Rico.” (P4, FG1)<br>“My daughter has friends whose fathers fish... sometimes they have a really hard time selling their seafood.” (P2, FG1)<br>“If you have a polluted area and the fishermen can no longer go out to fish...” (P7, FG1)<br>“...leaving Guánica because if there’s nothing here, people will just keep leaving the town.” (Fisherman, Interview 1) |
| 4.2 | Economic nostalgia | Memories of Guánica's economic prosperity during the era of petrochemical industries, agriculture, and tourism. | "Because before when I arrived to Guánica there were many businesses on the edge of the pier, right? There are hardly any businesses there. I think they have been affected." (P5, FG2)<br>"Yes, but as time went on, with the neglect of some things and others, the businesses closed down too." (P5, FG1) |
| 4.3 | Cost of living and financial struggle | Job market opportunities and financial limitations found locally. | "The family... is struggling... they left us with pollution and unemployment among the fishermen." (Fisherman, Interview 1)<br>"My son loves Guánica... but he can't live in Guánica because his job has to be in the metropolitan area, because there's nowhere to work here." (P2, FG1)<br>"We need the economy of this town to get ahead, because as I've already mentioned, it's one of the poorest towns in Puerto Rico." (P2, FG2)<br>"That's why we're the poorest town in Puerto Rico." (P8, FG1) |
| **5.0 Awareness and knowledge of PCBs** |  |  |  |
| 5.1 | Understanding of PCB contamination | Community level of understanding or information received about PCB contamination. | "What is PCB? I didn't know until today." (P4, FG2)<br>"Since the beginning, we knew there was pollution, because it wasn't just the contamination from chemicals and heavy metals that existed back then. There was also sewage pollution." (Fisherman, Interview 2) |
| 5.2 | Contamination information sources | Where people receive or discuss information concerning contamination (TV, radio, community events). | "They gave us the material... about the harm one can suffer... from inhaling that polluted air." (P4, FG1)<br>"Even the EPA has come here... they held meetings." (P4, FG1)<br>"Someone shared it with me... online." (P8, FG1)<br>"I get it from television... and from rumors I hear from people." (P7, FG1) |
| 5.3 | Communication gaps | Lack of accessible, consistent information. | "And it hasn't been widely publicized in the community so that people can attend these meetings and be informed in advance." (P1, FG2)<br>"I kept waiting to see if anything had been done, if it had been said that what was going to be done, or if it was already being done." (P3, FG1)<br>"I know this issue has been debated for<br>several years, right? And what's still going on? What stage of the plan are we at?" (P1, FG2) |
| **6.0 Trust and collaboration** |  |  |  |
| 6.1 | Perceptions of agencies and organizations | Mentions of agencies tasked with cleanup efforts in the area (EPA, municipality) and organizations bringing awareness, including trust or skepticism toward them. | "There is money, but they're squandering the money earmarked for combating pollution." (P3, FG1)<br>"You can't put goats in charge of lettuce." (P2, FG2)<br>"At first, I didn't trust any of this, because sometimes you think it's just another outsider coming in who's going to do something and then doesn't do anything." (Fisherman, Interview 2)<br>"And the negligence of the authorities in this regard is worrying. The struggle is immense... We must continue educating and we must continue fighting." (P3, FG2) |
| 6.2 | Accountability and corruption concerns | Beliefs that money or promises are misused. | "...the money allocated for combating pollution... is being squandered... there must be justice for those who are misappropriating the funds." (P1, FG2)<br>"...if I give someone ten pesos, they might steal them the first time, but they won't be able to steal them the second time." (P6, FG1) |
| 6.3 | Desire for transparent collaboration | Requests for updates, involvement, and clarity. | "I haven't seen any action other than... going to collect samples." (Fisherman, Interview 2)<br>"Well, let's hope this meeting we had doesn't just fizzle out." (P1, FG1)<br>"To give the community an update on what they found in Guánica Bay... to explain it a little better." (Fisherman, Interview 1)<br>"You don't see the EPA... holding federal meetings... We here only listen and get scared." (P4, FG1) |
| **7.0 Community voice and participation** |  |  |  |
| 7.1 | Involvement in decision-making | Level of community inclusion in discussions about the bay or cleanup. | "I agree, but right now we're also interested." (P3, FG2) |
| 7.2 | Barriers to participation | Lack of information, awareness, or logistics that limit involvement. | "...all I think is that this shouldn't just become a dead letter or a shelved letter... I haven't seen this being proactive." (P1, FG1) |
| 7.3 | Community | Responsibilities as citizens with | "...it's not all the fault of the governments |
|  | responsibility | respect to environmental actions. | as citizens; it's also a responsibility of individuals..." (P6, FG1) |
| 7.4 | Community learning and shared knowledge | How group conversations foster awareness and shared understanding of local issues. | "I'm enjoying this conversation because I'm learning things about Guánica that I didn't know." (P2, FG1) |
| **8.0 Resilience and adaptation** |  |  |  |
| 8.1 | Coping and acceptance | Descriptions of how the community continues to live despite the contamination. | "I continue to live peacefully; I am not afraid." (P5, FG1) |
| 8.2 | Hope for change | Optimism for a better future and continued advocacy. | "I have faith that something good will happen; this town doesn't give up." (P6, FG1)<br>"I hope that this time they actually do something and it doesn't just remain empty promises." (P3, FG2) |
| **9.0 Recommendations for action** |  |  |  |
| 9.1 | Environmental cleanup | Need for environmental cleaning and restoration processes to protect human health and ecosystems. | "Guánica... needs its bay cleaned up... the necessary cleanups to clear the areas so more tourists can come." (P2, FG1)<br>"I don't know if there's a way to clean the bay itself, to get rid of PCBs or other metals... If there is, they should implement it here." (P6, FG1) |
| 9.2 | Public health initiatives | Public health efforts targeting Guánica's population. | "We put up a sign on the beach that said, 'Remember you took your cell phone, your dog, your child, take your trash with you as well.'" (P1, FG1)<br>"I'd like the Health Department to do some work... How are all the people doing so they can check everything, even our bodies. They should give us some tests." (P1, FG1) |
| 9.3 | Community education and youth engagement | Teaching future generations about health and the environment. | "We need to raise awareness among young people because it's truly sad... If they don't teach their children from a young age to keep things clean, what's so hard about taking the trash in the car and throwing it away when they get home?" (P1, FG1)<br>"I know that if you guys say, like, 'Oh, let's do a workshop or a seminar,' we'll explain the information, and then we'd be grateful if you spread the word. That would work." (P7, FG2) |
| 9.4 | Economic and tourism revitalization | Linking cleanup to job creation and tourism. | "...an economic recovery... so that it can move forward... many young people have left... we are mostly old people." (P1, FG1) |

**Table A2.**
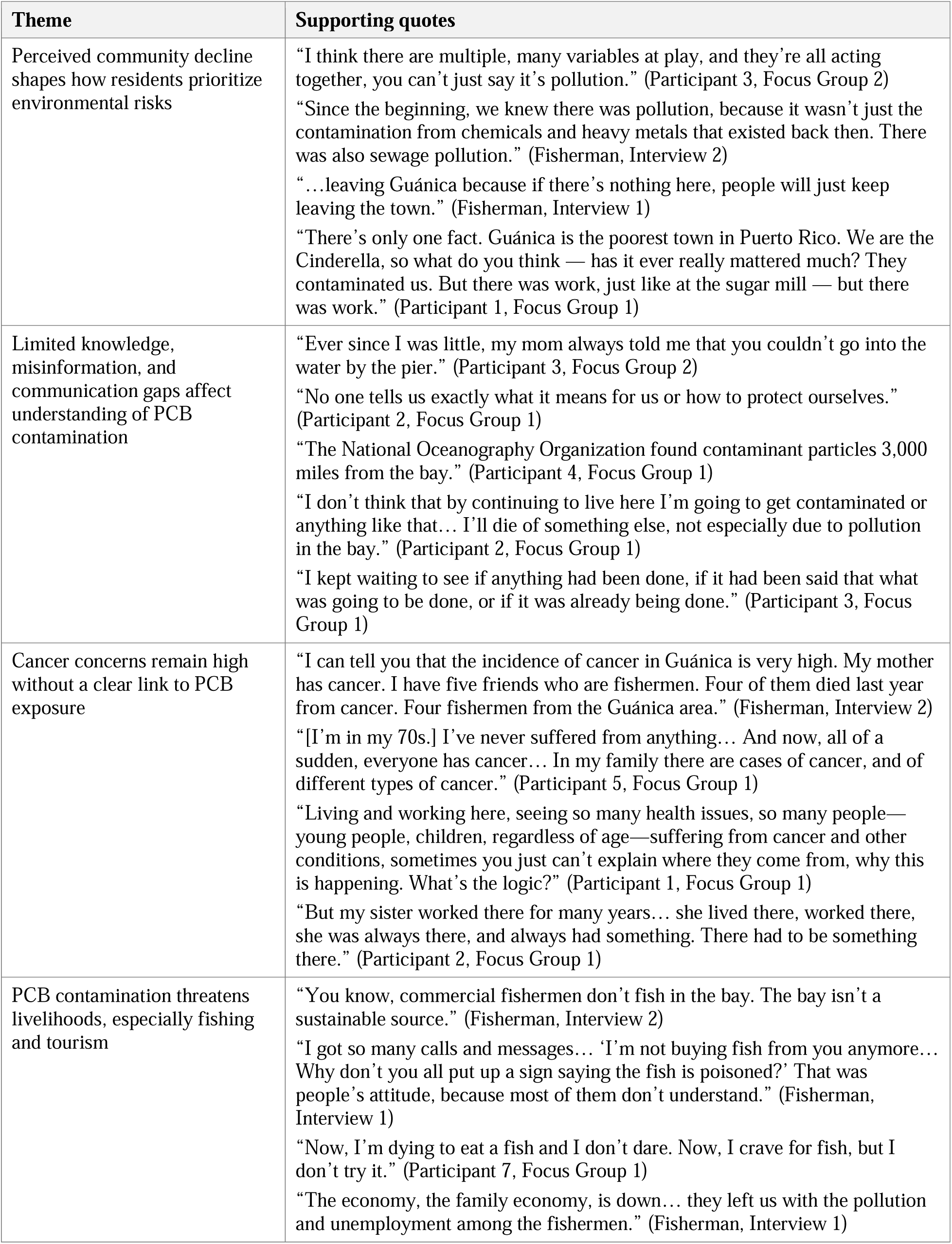
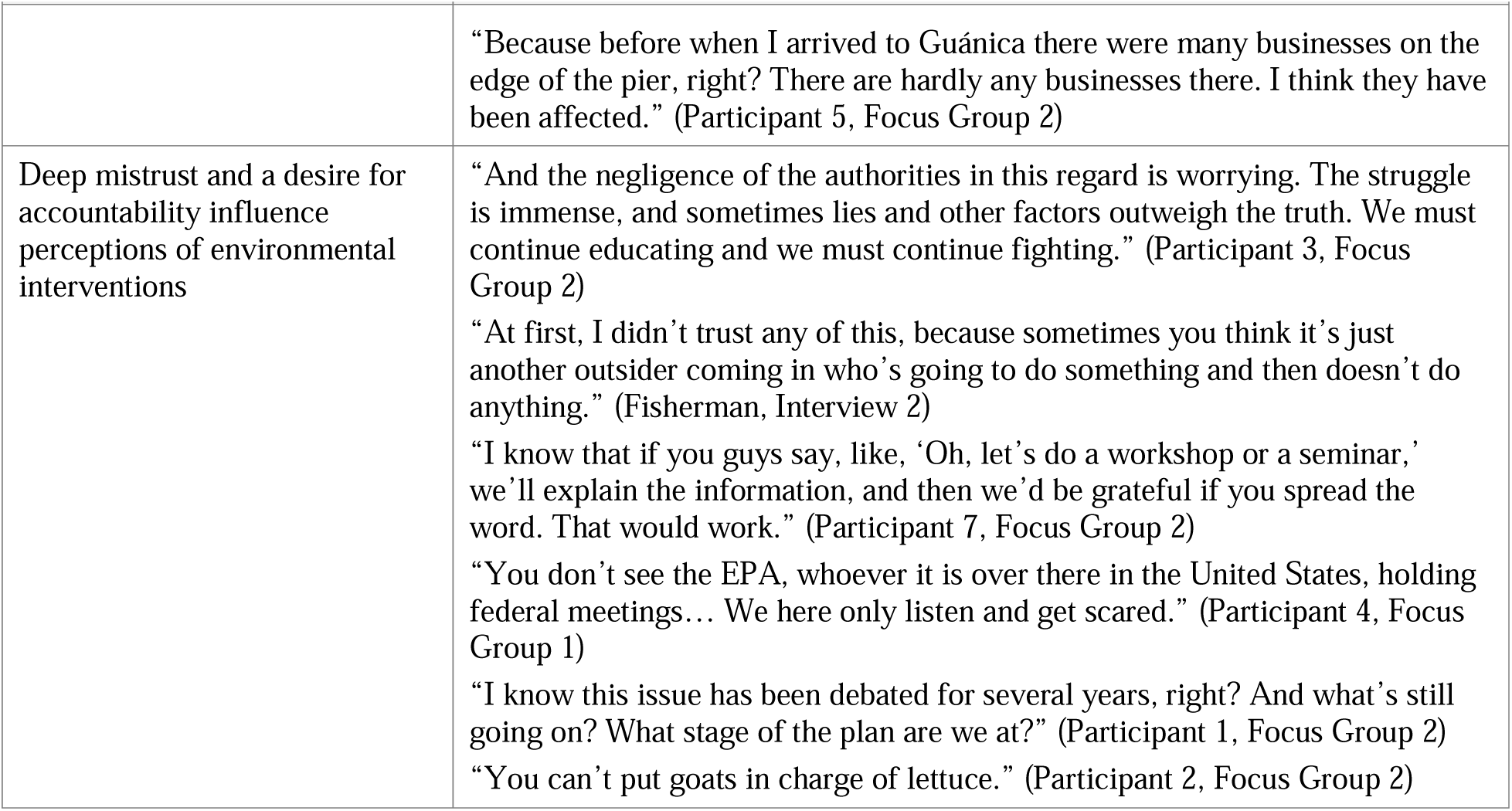
Qualitative themes and supporting quotes.

| Theme | Supporting quotes |
| --- | --- |
| Perceived community decline shapes how residents prioritize environmental risks | “I think there are multiple, many variables at play, and they’re all acting together, you can’t just say it’s pollution.” (Participant 3, Focus Group 2)<br>“Since the beginning, we knew there was pollution, because it wasn’t just the contamination from chemicals and heavy metals that existed back then. There was also sewage pollution.” (Fisherman, Interview 2)<br>“...leaving Guánica because if there’s nothing here, people will just keep leaving the town.” (Fisherman, Interview 1)<br>“There’s only one fact. Guánica is the poorest town in Puerto Rico. We are the Cinderella, so what do you think — has it ever really mattered much? They contaminated us. But there was work, just like at the sugar mill — but there was work.” (Participant 1, Focus Group 1) |
| Limited knowledge, misinformation, and communication gaps affect understanding of PCB contamination | “Ever since I was little, my mom always told me that you couldn’t go into the water by the pier.” (Participant 3, Focus Group 2)<br>“No one tells us exactly what it means for us or how to protect ourselves.” (Participant 2, Focus Group 1)<br>“The National Oceanography Organization found contaminant particles 3,000 miles from the bay.” (Participant 4, Focus Group 1)<br>“I don’t think that by continuing to live here I’m going to get contaminated or anything like that... I’ll die of something else, not especially due to pollution in the bay.” (Participant 2, Focus Group 1)<br>“I kept waiting to see if anything had been done, if it had been said that what was going to be done, or if it was already being done.” (Participant 3, Focus Group 1) |
| Cancer concerns remain high without a clear link to PCB exposure | “I can tell you that the incidence of cancer in Guánica is very high. My mother has cancer. I have five friends who are fishermen. Four of them died last year from cancer. Four fishermen from the Guánica area.” (Fisherman, Interview 2)<br>“[I’m in my 70s.] I’ve never suffered from anything... And now, all of a sudden, everyone has cancer... In my family there are cases of cancer, and of different types of cancer.” (Participant 5, Focus Group 1)<br>“Living and working here, seeing so many health issues, so many people— young people, children, regardless of age—suffering from cancer and other conditions, sometimes you just can’t explain where they come from, why this is happening. What’s the logic?” (Participant 1, Focus Group 1)<br>“But my sister worked there for many years... she lived there, worked there, she was always there, and always had something. There had to be something there.” (Participant 2, Focus Group 1) |
| PCB contamination threatens livelihoods, especially fishing and tourism | “You know, commercial fishermen don’t fish in the bay. The bay isn’t a sustainable source.” (Fisherman, Interview 2)<br>“I got so many calls and messages... ‘I’m not buying fish from you anymore... Why don’t you all put up a sign saying the fish is poisoned?’ That was people’s attitude, because most of them don’t understand.” (Fisherman, Interview 1)<br>“Now, I’m dying to eat a fish and I don’t dare. Now, I crave for fish, but I don’t try it.” (Participant 7, Focus Group 1)<br>“The economy, the family economy, is down... they left us with the pollution and unemployment among the fishermen.” (Fisherman, Interview 1) |
|  | “Because before when I arrived to Guánica there were many businesses on the edge of the pier, right? There are hardly any businesses there. I think they have been affected.” (Participant 5, Focus Group 2) |
| Deep mistrust and a desire for accountability influence perceptions of environmental interventions | “And the negligence of the authorities in this regard is worrying. The struggle is immense, and sometimes lies and other factors outweigh the truth. We must continue educating and we must continue fighting.” (Participant 3, Focus Group 2)<br>“At first, I didn’t trust any of this, because sometimes you think it’s just another outsider coming in who’s going to do something and then doesn’t do anything.” (Fisherman, Interview 2)<br>“I know that if you guys say, like, ‘Oh, let’s do a workshop or a seminar,’ we’ll explain the information, and then we’d be grateful if you spread the word. That would work.” (Participant 7, Focus Group 2)<br>“You don’t see the EPA, whoever it is over there in the United States, holding federal meetings... We here only listen and get scared.” (Participant 4, Focus Group 1)<br>“I know this issue has been debated for several years, right? And what’s still going on? What stage of the plan are we at?” (Participant 1, Focus Group 2)<br>“You can’t put goats in charge of lettuce.” (Participant 2, Focus Group 2) |

